# Opportunities and challenges when using record linkage of routinely collected electronic health care data to evaluate outcomes of systemic anti-cancer treatment in clinical practice

**DOI:** 10.1101/2021.05.28.21257611

**Authors:** Tanja Mueller, Jennifer Laskey, Kelly Baillie, Julie Clarke, Christine Crearie, Kimberley Kavanagh, Janet Graham, Kathryn Graham, Ashita Waterson, Robert Jones, Amanj Kurdi, David Morrison, Marion Bennie

## Abstract

**Objectives:** To discuss the opportunities and challenges when applying an electronic record linkage methodology with respect to systemic anti-cancer therapy, and to highlight some of the potential pitfalls spanning the entire breadth and depth of the research process.

**Design:** Retrospective cohort studies using routinely collected, administrative health data. Setting: Scotland

**Results:** Studies conducted to-date have indicated that record linkage of routinely collected data to determine outcomes of treatment with cancer medicines is feasible, albeit currently within certain limits. While the general description of patient populations and the calculation of median overall survival are well supported, prevailing issues with combining data across regional boundaries and the limited availability of some variables (including molecular pathology data and information regarding toxicities) may restrict the extent of analyses feasible.

**Conclusion:** There is scope to conduct large cohort studies to generate results from clinical practice using linkage of routinely collected health care data within a reasonable time frame; however, close collaboration between researchers, data controllers, and clinicians is required in order to obtain valid and meaningful results.

**Article Summary:** *Strengths and limitations of this study:*

- This is the first description of a comprehensive programme aimed at evaluating the feasibility of linking routinely collected health data to evaluate the outcomes of systemic anti-cancer treatment in clinical practice.
- The multi-disciplinary study team comprised both academics and clinicians, with a wide range of skills, expertise, and experience.
- While a number of studies focusing on different cancers have successfully been conducted, not all research questions of interest could be fully answered due to some existing data gaps.
- Nevertheless, study findings have been discussed with health care professionals across Scotland, and have started to have an impact on shared patient-clinical decision making.

## INTRODUCTION

Globally, medicines account for a significant proportion of healthcare interventions and resource use.[1] In the setting of cancer, systemic anti-cancer treatments (SACT) are used at different stages in the treatment pathway – increasingly on a continuous basis until disease progression rather than as a fixed course of treatment. With a general trend towards aging populations,[2,3] improvements in the early detection of cancer, and the availability of advanced treatment options, many more patients are now living with cancer, and are being treated with some form of SACT for a significant proportion of their cancer journey.[4,5]

Evidence to support the safety and efficacy of new cancer medicines is obtained through randomised clinical trials (RCTs), which is then used to support licencing applications. However, results based on RCTs – the gold standard with regards to evaluating the effect of drugs on patients – may not always translate into similar benefits in routine clinical practice, with absolute outcomes such as survival frequently being inferior in a real world setting compared to those reported in the pivotal trials.[6,7] Moreover, side effects from medicines, which can be particularly prevalent with SACT, may occur with increased frequency in populations encountered in routine healthcare settings. There are a number of potential reasons for inferior outcomes being observed in routine clinical practice; most importantly, participants included in RCTs are often not fully representative of patient populations, having been carefully selected based on pre-specified inclusion and exclusion criteria. Patients receiving these medicines in routine settings are much more varied – generally being older and frailer than trial participants, with more co-morbidities.[8–10]

The discrepancy between clinical trial results and treatment outcomes observed in clinical practice poses a risk-benefit challenge for prescribers and patients, especially in the context of SACT which often has modest survival gains and may negatively impact quality of life. Hence, in order to ensure that cancer treatments are of benefit to patients, post-marketing studies evaluating the clinical effectiveness and safety of these medicines in routine practice – as opposed to the tightly controlled settings of RCTs – are essential. Such observational studies are, by their nature, not able to measure the incremental benefits associated with the medicine, as it is rarely possible to derive an appropriate control population; however, they can help clinicians, patients, and policymakers better understand the likely outcomes among patients treated in clinical practice.

With recent advances in digital infrastructure and the advent of electronically available health records, linkage of routinely collected administrative health data has emerged as a promising evaluation method for observational studies – mainly because record linkage offers the opportunity to conduct large-scale cohort studies, while simultaneously enabling the inclusion of information from much broader sources of data than was previously feasible.[11] Nevertheless, using administrative health data is not without its challenges; although technical difficulties have largely been overcome, certain issues – mainly regarding privacy and data availability/accessibility – remain topical.[12,13] Furthermore, cancer treatment is very complex, not least due to the different purposes of treatment – ranging from neo-adjuvant intent, i.e. treatment given as a first step in order to reduce the size of a tumour prior to the main treatment (such as surgery or radiotherapy), to palliative treatment, aimed at alleviating symptoms in patients with incurable disease.

### Key requirements for observational studies assessing outcomes of cancer medicines

Crucial aspects to be considered when conducting studies evaluating SACT treatment outcomes in clinical practice fall into three major categories:

- Data – demographic and clinical information to allow patients, treatments, and outcomes to be described;
- Information governance – permissions to process data; and
- Interdisciplinary collaboration – the strategic and operational groups enabling and advising the work.

Data requirements mainly relate to the four areas of patient identification, treatment exposure, patient baseline characteristics, and treatment outcomes. While death records and hospitalisation data are vital to facilitate the assessment of treatment outcomes, complete regimen and indication data for the population are primary pre-requisites for any study assessing SACT treatment – allowing the identification of study eligible patients and the reliable definition of the exposure of interest, thereby decreasing the risk of bias. Summarising patient baseline characteristics such as age or comorbidities is necessary to support the interpretation of study findings, and to enable comparisons to trial populations or other published studies; for example, several disease-related factors are recognised to be important confounders and/or predictors of treatment effectiveness and/or survival, including cancer staging and the Eastern Cooperative Oncology Group Performance Status (ECOG PS).[14]

Proper information governance demands that data are processed according to relevant laws and guidance, which in Europe includes, for instance, the General Data Protection Regulation. Formal ethics committee approval may not be required if evaluation of outcomes is considered to be part of clinical audits – a component of good clinical practice – but the specifics of implemented procedures may differ depending on context. There is usually a requirement to highlight the potential benefit of a proposed study to patients and/or a health care system, as well as provisions relating to data anonymisation, researcher training, and the safety and security of work environments and associated IT infrastructure.

Considering the complexity of the topic and the amount and nature of data required to conduct studies focusing on outcomes of cancer medicines in clinical practice, interdisciplinarity of the study team and broad academic and health services support is essential; clinical expertise, especially in assessing patients for SACT and SACT prescribing, is crucial. Furthermore, governmental/health system engagement is recommended to facilitate rapid translation of findings into clinical practice.

### The Cancer Medicines Outcomes Programme

Following publication of the “Cancer Action Plan” and in line with the national Digital Strategy, [15,16] in 2016, the Scottish Government made funding available for the Cancer Medicines Outcomes Programme (CMOP), a collaborative project aimed at evaluating the feasibility of using routinely collected electronic health data to determine outcomes of SACT in clinical practice in Scotland. The programme team comprises academics, quantitative and qualitative researchers, and clinicians – spanning a variety of disciplines, specialties, and areas of expertise. The main objective of the initial three-year programme was to set up a series of incremental studies to test existing linkage capabilities and assess data quality with regards to cancer treatment outcomes in local populations.

The purpose of this paper is to discuss the opportunities and challenges when applying an electronic record linkage methodology with respect to SACT by showcasing a series of CMOP studies conducted between 2017 and 2020. Although firmly situated within a Scottish setting, the majority of challenges encountered are likely not exclusive to Scotland.

## METHODS

### Study context

Scotland has a population of approximately 5.4 million, and all residents are covered by the National Health Services (NHS) Scotland – a tax-based system offering healthcare free of charge at the point of delivery, including hospital care and prescription medicines. Cancer services are provided through three regional cancer networks: the West of Scotland Cancer Network (WoSCAN), covering four Health Boards including NHS Greater Glasgow & Clyde (NHS GGC), the largest Health Board in Scotland (Health Boards are regional organisations responsible for all health care provisioning to their respective populations); the South-East Cancer Network (SCAN), which covers four Health Boards and includes the capital city, Edinburgh; and the North Cancer Alliance (NCA), comprising six Health Boards spread out across the Scottish Highlands and Islands, mostly covering large geographical areas but relatively small populations.[17–19] Due to close collaboration with NHS GGC, initial CMOP projects, serving as pilot studies, focused on NHS GGC, which covers approximately 1.2 million people (or 20% of the overall Scottish population); subsequent projects were intended to test the scalability of the methodology and aimed to include all patients residing within WoSCAN, covering almost 50% of the total population of Scotland.[3]

### Data sources

In Scotland, a range of routinely collected, national-level administrative datasets are available for research purposes, ranging from medicines dispensed in primary care to death records. In addition, clinical datasets providing radiotherapy treatment information and laboratory test results, curated by individual Health Boards, can be made available. Details of the main datasets used for CMOP studies are described in table 1.

**Table 1:** Main data sources used for CMOP record linkage projects

| Data source | Content | Data controller |
| --- | --- | --- |
| National Register of Scotland (NRS) [54] | Registration of life events – death records (date and cause of death, ICD10 codes) | Public Health Scotland |
| Scottish Cancer Registry (SMR06) [54] | Collection of incident cancer registrations from across the health system; date of diagnosis, ICD10 and ICD-O codes, and details relating to the tumour | Public Health Scotland |
| Scottish Morbidity Records, outpatient attendances (SMR00) [54] | Episode level data on outpatient clinic attendances, including diagnostic codes and procedures undertaken | Public Health Scotland |
| Scottish Morbidity Records, inpatient and day case dataset (SMR01) [54] | Episode level data on acute hospital admissions, including diagnostic codes (ICD10) and procedures undertaken (OPCS4 codes) | Public Health Scotland |
| Prescribing Information System (PIS) [54] | Primary care prescribing, including date of prescription and dispensing, and drug name, dose, and quantity | Public Health Scotland |
| Chemotherapy Electronic Prescribing and Administration System (CEPAS) [20] | Systemic anti-cancer therapy prescribing, including diagnosis, drug, dose, indication, date of administration, and cycle number | Health Board |
| Scottish Care Information (SCI store) [55] | Laboratory test results (biochemistry and haematology); comprising test name, date, and value | Health Board |
| ARIA | Radiotherapy records, including indication (ICD10 codes), dose/fraction, and dates of administration | Health Board |

ICD10 – International Classification of Diseases, 10<sup>th</sup> edition; ICD-O – International Classification of Diseases for Oncology; OPCS4 – (Office of Population Censuses and Surveys) Classification of Interventions and Procedures

### Study design

All CMOP studies conducted to-date (as listed in table 2) have been designed as retrospective cohort studies. For practical reasons and to ensure that study cohorts were composed appropriately, patients were identified based on cancer diagnosis and/or treatment, within a defined time frame, through the Chemotherapy Electronic Prescribing and Administration System (CEPAS), a system used to record the prescription of SACT.[20] Record linkage with other relevant datasets has been enabled through the availability of a unique patient identifier in Scotland, the Community Health Index (CHI) number; CHI numbers are assigned to every resident (at birth or entry into the Health Care System), and their use is mandatory across the Health and Social Care system in Scotland.[21] While all studies had a very similar design, analytical methods were adapted to fit specific questions depending on cancer type.

**Table 2:**
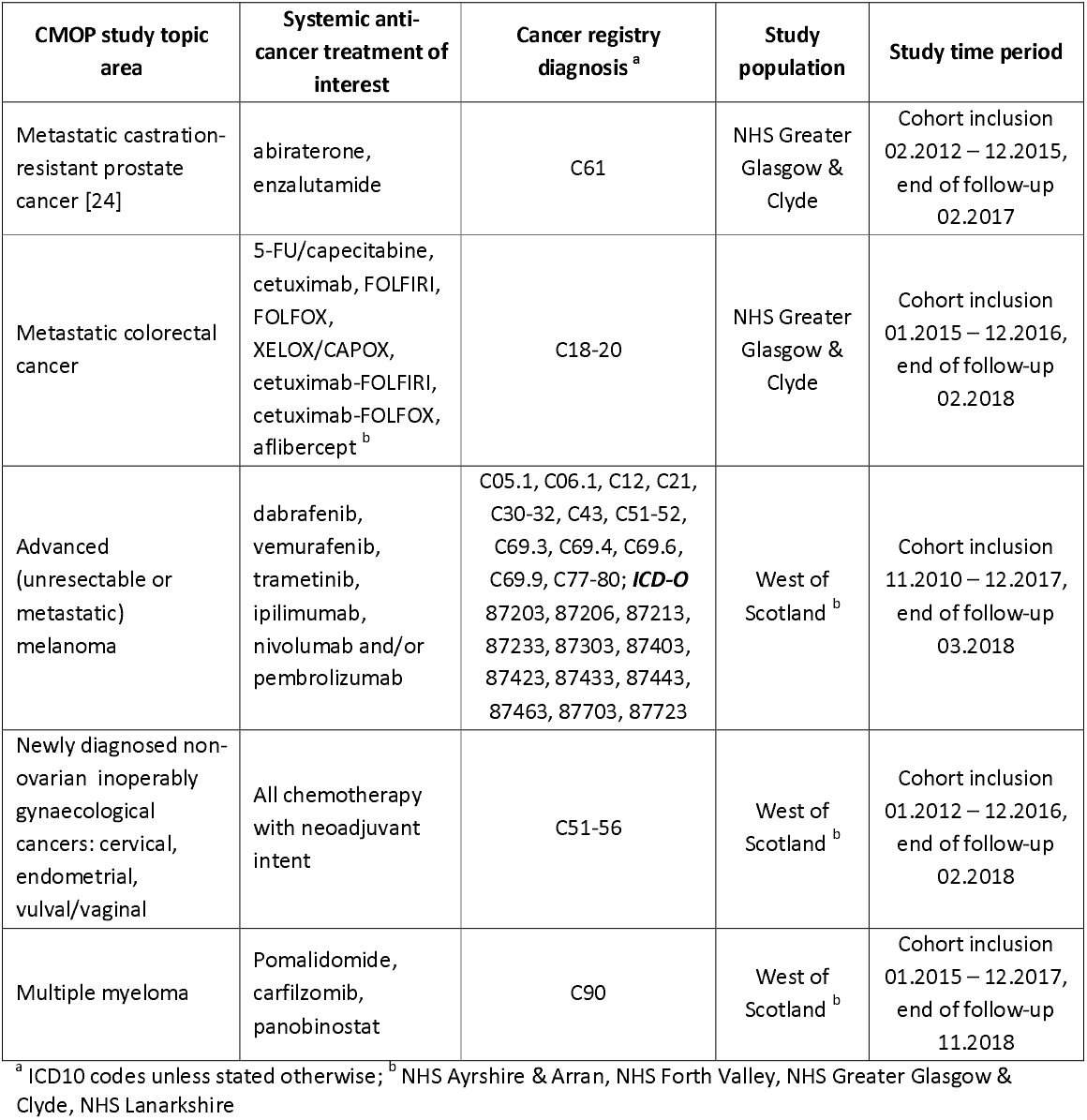
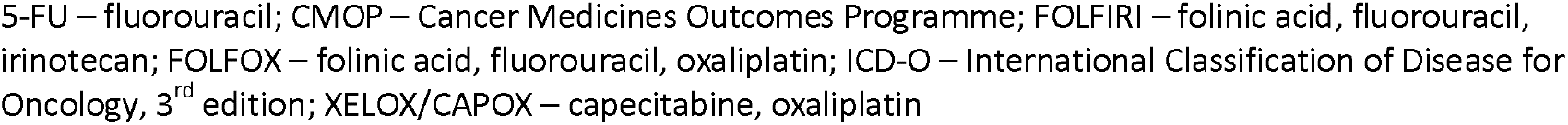
Cancer Medicines Outcomes Programme studies completed or in-progress, 2017 – 2020

| CMOP study topic area | Systemic anti-cancer treatment of interest | Cancer registry diagnosis <sup>a</sup> | Study population | Study time period |
| --- | --- | --- | --- | --- |
| Metastatic castration-resistant prostate cancer [24] | abiraterone, enzalutamide | C61 | NHS Greater Glasgow & Clyde | Cohort inclusion 02.2012 – 12.2015, end of follow-up 02.2017 |
| Metastatic colorectal cancer | 5-FU/capecitabine, cetuximab, FOLFIRI, FOLFOX, XELOX/CAPOX, cetuximab-FOLFIRI, cetuximab-FOLFOX, aflibercept <sup>b</sup> | C18-20 | NHS Greater Glasgow & Clyde | Cohort inclusion 01.2015 – 12.2016, end of follow-up 02.2018 |
| Advanced (unresectable or metastatic) melanoma | dabrafenib, vemurafenib, trametinib, ipilimumab, nivolumab and/or pembrolizumab | C05.1, C06.1, C12, C21, C30-32, C43, C51-52, C69.3, C69.4, C69.6, C69.9, C77-80; **ICD-O** 87203, 87206, 87213, 87233, 87303, 87403, 87423, 87433, 87443, 87463, 87703, 87723 | West of Scotland <sup>b</sup> | Cohort inclusion 11.2010 – 12.2017, end of follow-up 03.2018 |
| Newly diagnosed non-ovarian inoperably gynaecological cancers: cervical, endometrial, vulval/vaginal | All chemotherapy with neoadjuvant intent | C51-56 | West of Scotland <sup>b</sup> | Cohort inclusion 01.2012 – 12.2016, end of follow-up 02.2018 |
| Multiple myeloma | Pomalidomide, carfilzomib, panobinostat | C90 | West of Scotland <sup>b</sup> | Cohort inclusion 01.2015 – 12.2017, end of follow-up 11.2018 |

<sup>a</sup> ICD10 codes unless stated otherwise; <sup>b</sup> NHS Ayrshire & Arran, NHS Forth Valley, NHS Greater Glasgow & Clyde, NHS Lanarkshire
5-FU – fluorouracil; CMOP – Cancer Medicines Outcomes Programme; FOLFIRI – folinic acid, fluorouracil, irinotecan; FOLFOX – folinic acid, fluorouracil, oxaliplatin; ICD-O – International Classification of Disease for Oncology, 3<sup>rd</sup> edition; XELOX/CAPOX – capecitabine, oxaliplatin

### Data management

Permission to use all requested datasets for CMOP studies has been approved by the Public Benefit and Privacy Panel for Health and Social Care,[22] and access has been granted through the Glasgow Safe Haven – a secure, closed environment hosted by the University of Glasgow and supported by NHS GGC.[23] Data has been pseudonymised, and no identifying information has been made available to researchers; results to be released from the Safe Haven are subject to statistical disclosure procedures. Therefore, additional ethical approval was not required.

### Patient and Public Involvement

While patients or the public were not directly involved in the design, conduct, or dissemination of individual studies, the CMOP Board – responsible for the general oversight of the project – includes patient advocacy representatives.

## FINDINGS

### Identifying patients

For the majority of CMOP studies, the study populations were initially identified through CEPAS, where patients are assigned both an indication for treatment (the diagnosis) and a regimen (the treatment), accompanied by a treatment intent (e.g. adjuvant or palliative); the Cancer Registry (SMR06), while being used to confirm diagnoses, did not provide sufficient information to be used for cohort identification. In the first CMOP study for example, focusing on patients with metastatic castration-resistant prostate cancer (mCRPC),[24] patients were identified based on the treatment prescribed on CEPAS (abiraterone or enzalutamide), and the indication was confirmed by the attached mCRPC diagnoses in CEPAS as well as the patients’ diagnoses of prostate cancer as recorded in SMR06. Similarly, patients for a study intended to analyse SACT outcomes in patients with metastatic melanoma were identified within CEPAS through a combination of diagnosis and treatment regimen, and eligibility for study inclusion was confirmed via ICD10 codes as recorded in SMR06 (see also table 2 for further details).

In contrast, identification of patients with some other cancers was more challenging. For instance, the identification of patients newly diagnosed with a range of gynaecological cancers (cervical, endometrial, and vaginal/vulval cancer) subject to neo-adjuvant treatment required a considerable amount of clinical input, for two main reasons: first, neo-adjuvant therapy was not easily identifiable in CEPAS for some cancer types, as this sub type was not necessarily included within the prescribing setup (where diagnoses can be selected from a drop-down menu); and second, the complex nature of these cancers impacts the reliability and consistency of the data available, since diagnoses may evolve as more information becomes available over time (e.g. from biopsies or scans). Consequently, identifying patients comprised several steps and required information from a number of datasets in order to exclude study ineligible patients (e.g. those with recurrent disease), including: details of received SACT; timing between diagnosis and initiation of SACT; and previous/concurrent treatment for the cancer of interest. The flowchart in figure 1 illustrates the complexity of the patient identification process.

**FIGURE 1:**
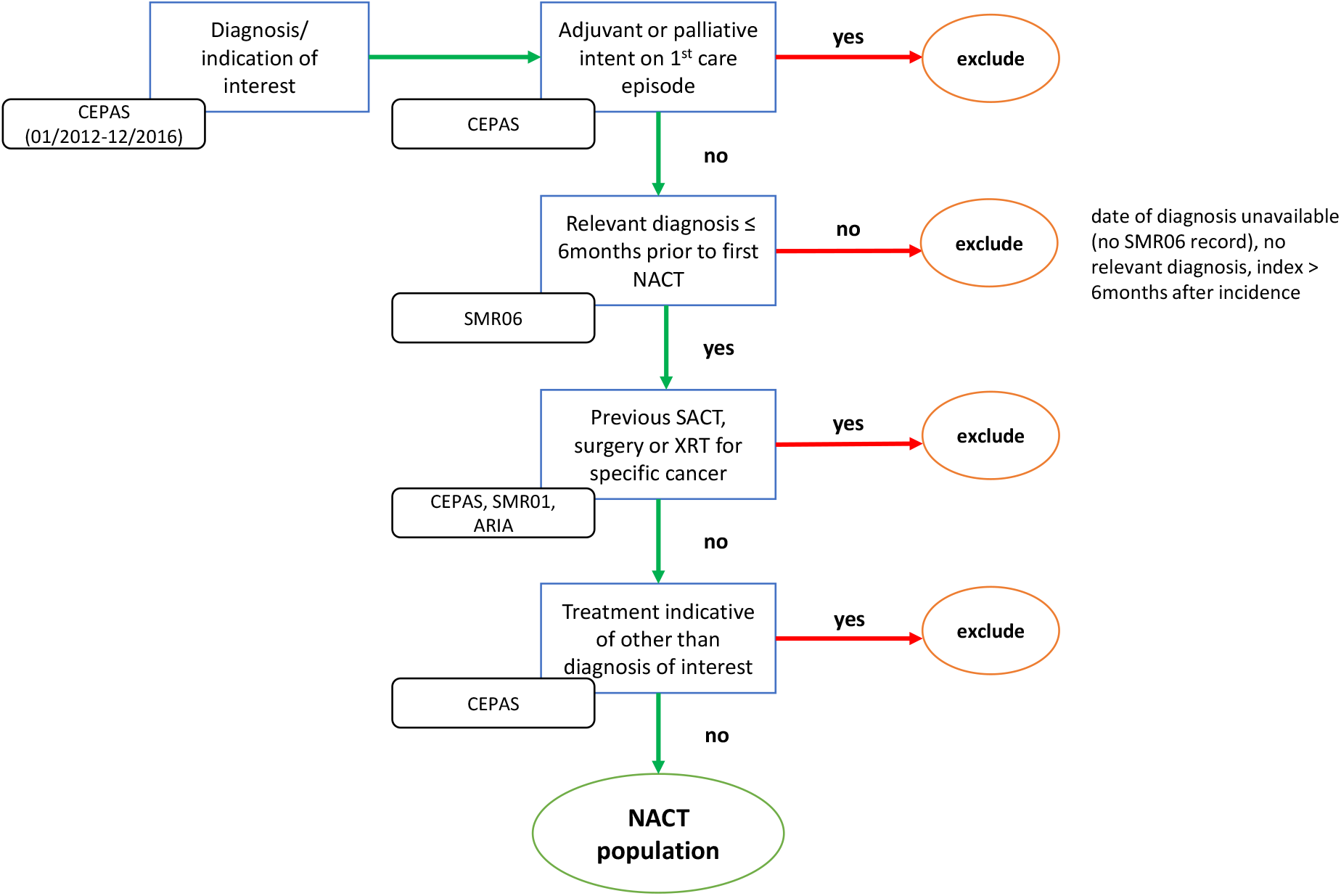
Generic flowchart detailing identification of a neo-adjuvant chemotherapy patient cohort in gynaecological cancers. ARIA – radiotherapy treatment records dataset; CEPAS – Chemotherapy Prescribing and Administration System; NACT – neo-adjuvant chemotherapy; SACT – systemic anti-cancer treatment; SMR01 – Scottish Morbidity Records, inpatient dataset; SMR06 – Scottish Cancer Registry; XRT – radiotherapy

### Defining exposure

Exposure to cancer medicines is comprehensively collected within CEPAS, including the name of the regimen, the drugs given along with their prescribed dose, and dates of administration. While defining exposure can be straightforward – as, for instance, in the case of mCRPC patients were the treatments of interest (abiraterone and enzalutamide) were both single agent oral drugs given at a fixed dose – other therapies may require further interpretation of records to enable reliable categorisation (e.g. due to complex treatment schedules involving multiple drugs with some variability of sequencing and/or dosing of medication).

As an example: patients with metastatic colorectal cancer (mCRC) may be treated with a range of different regimens depending on circumstances, comprising both chemotherapy (most prominently fluorouracil (5-FU) and/or oxaliplatin) and/or targeted treatments (e.g. aflibercept or cetuximab). If applicable and appropriate, doublet or triplet therapy (combining more than one drug) is preferred over monotherapy. The number and combination of drugs given might, however, change over time for various reasons; changes may include “stepping up” of treatment, i.e. adding a drug to the previously prescribed drug – this might occur if there are initial concerns with regards to potential side effects and a patient’s fitness; or “stepping down” of treatment, i.e. removing a drug from an originally given regimen, usually due to a patient experiencing side effects relating to one of the drugs included in the combination.

Unfortunately, there is some variability in how these cases are handled in CEPAS, and it was not always possible to determine the reason for changes in treatment. Following in-depth discussions with clinicians involved in treating mCRC patients within NHS GGC, a set of rules was developed to facilitate the simplification of exposure while acknowledging the prescribing intention; these rules were based on a) the sequence of drugs prescribed; b) the number of cycles given; and c) the timing of different regimens and/or drugs. Figure 2 illustrates two examples.

**FIGURE 2:**
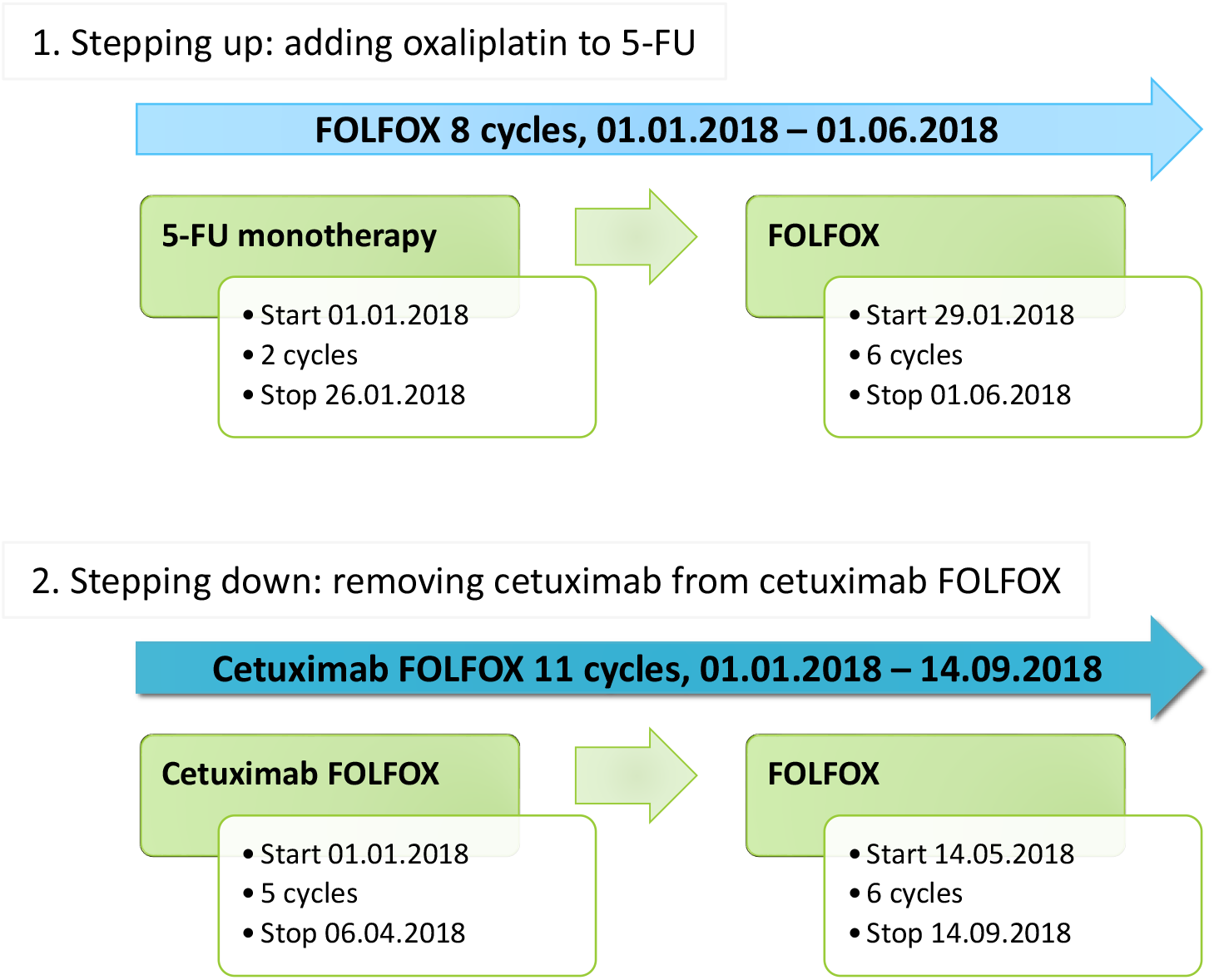
Examples of re-coding exposure due to stepping up or down of treatment. 5-FU – 5-fluoro-uracil; FOLFOX – folinic acid, fluorouracil, oxaliplatin

### Describing patient baseline characteristics

In Scotland, data on age and sex are incorporated into a person’s CHI number, and are included in all datasets. In addition, a number of geographical and socio-demographic information is available from the majority of health-related datasets; these usually include the Health Board of residence and the postcode area; an urban/rural classification;[25] and the Scottish Index of Multiple Deprivation (SIMD).[26] Information regarding co-morbidities can be obtained from hospital and outpatient clinic records, and data on medicines prescribed and dispensed in primary care may be used as a proxy for long-term conditions that do not require hospital admission; however, comorbidities might be missed if an individual has not been hospitalised and medication cannot be easily used as a proxy for the condition in question. Hospital records also comprise details with regards to procedures undertaken (see also table 1).

Regrettably, additional information relevant to cancer treatment is not always available in Scotland for record linkage studies for a number of reasons, with potential implications on the accuracy and comparability of results. Data stemming from (molecular) pathology – such as BRAF or RAS status – for instance are recorded locally by the departments conducting these tests, and stored in non-standardised formats (e.g. spreadsheets); while the presence of CHI numbers theoretically enables these datasets to be linked to other data sources, locating, accessing, and processing these files requires considerable resources, both in terms of time and technical expertise. This particularly affects studies aimed at combining patients across Health Boards, as resources and/or processes may differ considerably between regions. The Cancer Registry, in contrast, routinely captures patients from across Scotland; however, completion of all fields is not mandatory. While the registry offers the opportunity to collect additional useful information related to the disease including pathology/cell type and cancer staging based on the TNM (Tumour-Node-Metastases) system [27] or cancer-type appropriate methods (e.g. FIGO stage in patients with gynaecological cancers [28]), some of this data is frequently incomplete. In addition, records are entered at time of diagnosis and are usually not updated at recurrence – i.e. the location of metastases, for example, is only recorded if these were present at the time of diagnosis, but not if patients subsequently develop metastases. Furthermore, fields such as ECOG PS were non-mandatory within CEPAS in WoSCAN prior to July 2015, leading to incomplete data for earlier study cohorts.

### Defining treatment outcomes

The most common outcome measures as reported from both clinical trials and observational studies of cancer medicines are median overall survival (OS) and landmark survival figures (e.g. % alive after five years). Death records, comprising both date and cause of death, can easily be linked to other patient records in Scotland; therefore, analyses of these outcomes among cancer patients are well supported. CMOP studies conducted thus far have provided insights into OS among mCRPC patients treated with abiraterone or enzalutamide;[24] patients with metastatic melanoma or mCRC undergoing treatment with chemotherapy, targeted treatment, and/or immunotherapies; and patients with gynaecological cancers (cervical, endometrial, and vulval cancer) subject to neo-adjuvant treatment.

Nevertheless, progression-free survival (PFS) is increasingly used to establish early efficacy in advanced cancers in RCTs with small sample sizes and/or short duration.[29,30] PFS for solid tumours is usually based on imaging data; however, in haematological malignancies such as multiple myeloma, it is determined from changes in blood plasma levels of, e.g., paraprotein.[31] Laboratory test values are, to a certain extent, available for record linkage in Scotland; however, these are collected and stored locally within individual NHS Health Boards, and the existing infrastructure does not easily support the combination of data from different areas across Scotland for various reasons (including, but not limited to, the use of different lab equipment with diverging measurement standards and variability in the coding of tests). This, in effect, currently limits our capability of conducting studies across regions where laboratory test data is required for analyses. Imaging data – and, in extension, any other data not provided in a standardised format (such as patient-reported outcomes (PROMs) or free text information) – is not routinely available for use in record linkage studies. National Language Processing and other Machine Learning techniques enabling analyses of these data are, however, in development, and have in certain areas already proven their usefulness;[32,33] with further progress being made, PFS might be added to the list of treatment outcomes readily available for analyses in the future.

## DISCUSSION

Observational post-marketing studies substantially add to the evidence base upon which treatment decisions are made; this is true for all clinical areas, but especially in cancer where medicines often have a narrow risk/benefit ratio and promising RCT findings might not entirely translate into benefits in routine practice.[7–9] Although existing disadvantages of observational studies need to be kept in mind – most prominently the potential for confounding and other biases – these studies can offer a useful overview of the effectiveness and safety of medicines in clinical practice.[34,35] Furthermore, they present an invaluable opportunity to understand treatment within the ever-changing clinical environments outwith RCTs; this is of particular importance in light of COVID-19, which has considerably impacted the way SACT is being provided to patients.[36]

CMOP studies conducted to-date have indicated that record linkage of routinely collected data to determine outcomes of treatment with cancer medicines is feasible, albeit currently within certain limits. These studies offer a range of opportunities: informing clinicians of likely treatment outcomes in their local population; enabling patients to make better informed treatment choices, particularly at the end of life; allowing local clinical guidelines to be tailored to support the safe delivery of SACT and reduce the risk of side effects; and supporting the assessment of the relative value of medicines in local populations. The usefulness of record linkage to conduct pharmacoepidemiological studies in the area of cancer is supported by experiences gained in other clinical areas where this method has been used for many years; studies have, for example, used Scottish data to determine the effectiveness and safety of cardiovascular drugs; evaluate the effects of in-utero exposure to antihypertensive drugs on neonatal outcomes; and calculate the cost burden of Clostridium difficile infections, to name just a few.[37–39]

Although linkage of electronic health records has also frequently been used to conduct epidemiological studies in cancer – e.g. analysing the association between atopic dermatitis and the risk to develop colorectal cancer; or the impact of previous non-melanoma skin cancer on the occurrence of further primary tumours [40,41] – there is little evidence to-date with regards to studies focusing specifically on cancer medicine outcomes. Notable exceptions include, e.g., studies analysing: outcomes of chemotherapy among patients with ovarian cancer in the Netherlands; factors influencing breast cancer chemotherapy in the United States; and outcomes of immunotherapy in patients with lung cancer, again in the United States.[42–44] It is, however, worth pointing out that studies conducted in the United States were limited to beneficiaries of specific health insurance schemes, possibly limiting the comparability of findings due to selection bias.

### Key challenges

While the description of patient populations and the calculation of median OS are supported by record linkage, potential pitfalls span the entire breadth and depth of the research process – from data acquisition to selecting variables for statistical models. The following issues are not necessarily restricted to cancer studies but deserve particular consideration in this context.

### Data

Some data that would be useful for analyses is currently not routinely available for record linkage; or cannot easily be obtained. Most importantly, there are difficulties collating laboratory and molecular pathology data from across Health Boards, potentially affecting the scope of studies. In addition, across Health Boards, different versions of some data sources (particularly CEPAS) are used, making comparability of data and mapping and merging of datasets from different regions challenging.

The non-availability of specific variables might influence the ability to answer certain research questions; information regarding some potential toxicities of medicines is, for instance, not routinely captured across Scotland. Particular attention needs to be paid when attempting to use information that is contained in non-mandatory data fields of a dataset, such as socio-economic data (including, e.g., ethnicity or marital status); this data has most likely a high degree of missing-ness. Unfortunately, the same is true for data pertaining to treatment modifications and/or side effects in CEPAS. Furthermore, there is a considerable lag time (currently approximately 12-18 months) in the collection of Cancer Registry data, meaning that very recent data will likely be unavailable.

Most of these issues are, however, not unique to Scotland. Challenges with incomplete or out-of-date information in Cancer Registry data has, e.g., also been reported from the United States,[45,46] whereas details relating to SACT and laboratory test results appear to be incomplete in the Netherlands;[47] previous research conducted in the United States also advised caution when using data based on laboratory tests – not least due to diverging data recording conventions between health service providers.[48] Calls to standardise data collection, coding, and storage, and to implement nation-wide infrastructure with possible re-use of data in mind, have been made for many years.[49–51]

For the majority of datasets regularly used for research purposes, metadata is readily available; it is advisable to consult these prior to making any decisions with respect to study data. Nevertheless, it is highly recommended to collaborate with clinicians familiar with, ideally, both the treatment area of interest and the local systems used to generate and collect data. Many of the issues encountered while conducting the aforementioned studies – spanning from difficulties identifying the correct patients to uncertainties with regards to the appropriate categorisation of treatment exposure – were resolvable by discussing clinical procedures and treatment details with oncologists and other health care professionals working within these clinical settings.

### Information Governance

In Scotland, the current approval process to access routinely collected electronic data is both complicated and time consuming. Permissions to use data have to be sought from the Public Benefit and Privacy Panel for Health and Social Care,[22] potentially followed by an application to use one of four existing local Safe Havens [52] depending on how data is intended to be accessed. Subsequently, extraction, anonymisation, sense checking, and uploading of data onto the selected Safe Haven to grant researchers access may require input from several parties, depending on the datasets in question. Overall, this process might take several months – this needs to be taken into account at the planning stage of a study.[53]

### Interdisciplinary collaboration

CMOP has highlighted the benefit of researchers and clinicians collaborating on ambitious projects aimed at generating results useful for clinical practice by harnessing the wealth of routinely collected data. Study findings have been shared with health care professionals both within and outside WoSCAN, and clinicians are beginning to use the data to inform discussions regarding treatment decisions with their patients. In addition, initial conversations have taken place with the Scottish Government, intended to potentially impact future decisions related to data collection, recording, and access. Activities are already ongoing with regards to improving the Scottish Cancer Registry (level of completeness); upgrading CEPAS to version 6 (with the possibility of unifying data collection across Scotland and adding further information); and – potentially – implementing a Scotland-wide laboratory resource.

### Next steps

The Scottish Government has provided additional funding, enabling CMOP to continue its work.[36] Building on the experiences of the first three years, the next phase is aimed at solving some of the existing challenges; upscaling select projects to cover Scotland-wide patient populations; and further developing the applied methodology. While emphasising a drive towards quality improvement in Scotland, CMOP will also investigate the utility of data stemming from clinical practice to inform medicines assessment and reimbursement decisions; and explore the integration of patient reported outcomes measures related to cancer medicines into the Health Care System.

## CONCLUSION

Based on the experiences made in Scotland to-date, using electronic record linkage to evaluate the clinical effectiveness and safety of cancer medicines in day-to-day practice is feasible, and may offer scope to conduct large cohort studies to generate results from routine clinical practice within a reasonable time frame. Nevertheless, large-scale collaboration between researchers, data controllers, clinicians, and policy makers is urgently required in order to further improve existing systems and processes.

## Data Availability

NHS data is confidential, and is only available upon request subject to approval by a Caldicott Guardian/the Public Benefit and Privacy Panel for Health and Social Care.

## Acknowledgements

The work of the Cancer Medicines Outcomes Programme is only possible because of the wealth of information collected by the NHS as part of routine clinical practice.

## Author contributions

JL and MB obtained the funding. TM, JL, KB, JC, KK, JG, KG, AW, RJ, AK and MB participated in the conception of the studies. TM, KB, JC and CC arranged access to data. TM, KB, JC and CC cleaned the data and conducted the analyses. KK supervised the statistical analyses. JL, KB, JC, JG, KG, AW and RJ provided clinical advice and interpreted results. DM and MB provided strategic advice and further contextualised findings. TM drafted the manuscript. All authors read, critically revised, and approved the final draft.

## Patient consent for publication

Not required.

## Ethics approval

The use of NHS data was approved by the local Caldicott Guardian (NHS Greater Glasgow & Clyde) and/or the Public Benefit and Privacy Panel for Health and Social Care (study numbers 1617-0371/1917-0371; 1819-0055). In addition, accessing data was approved by the NHS Greater Glasgow & Clyde Safe Haven (study numbers GSH/17/ON/003; GSH/18/ON/012; GSH/19/ON/002); permissions include ethical approval (REC numbers for devolved rights to approve projects: 12/WS/0142; 17/WS/0237). All studies have been conducted in accordance with information governance standards; no identifiable data was available to researchers.

## REFERENCES

1 OECD. Health at a Glance 2019: OECD Indicators. Paris: OECD 2019. doi:10.1787/4dd50c09-en2 United Nations. Ageing. United Nations: Global Issues. https://www.un.org/en/sections/issues-depth/ageing/ (accessed 15 Oct 2020).

2 National Records of Scotland. Mid-2019 Population Estimates Scotland. Statistics and Data. 2020. https://www.nrscotland.gov.uk/statistics-and-data/statistics/statistics-by-theme/population/population-estimates/mid-year-population-estimates/mid-2019 (accessed 15 Oct 2020).

3 Miller KD, Nogueira L, Mariotto AB, et al. Cancer treatment and survivorship statistics, 2019. CA Cancer J Clin 2019;69:363–85. doi:10.3322/caac.21565

4 Forster V. Surviving cancer: how big data is helping patients live longer, healthier lives. 2019. https://www.lshtm.ac.uk/research/research-action/features/surviving-cancer-how-big-data-helping-patients-live-longer (accessed 15 Oct 2020).

5 Naci H, Davis C, Savović J, et al. Design characteristics, risk of bias, and reporting of randomised controlled trials supporting approvals of cancer drugs by European Medicines Agency, 2014-16: cross sectional analysis. BMJ 2019;366. doi:10.1136/bmj.l5221

6 Di Maio M, Perrone F, Conte P. Real-World Evidence in Oncology: Opportunities and Limitations. Oncologist 2020;25:e746–52. doi:10.1634/theoncologist.2019-0647

7 Jin S, Pazdur R, Sridhara R. Re-Evaluating Eligibility Criteria for Oncology Clinical Trials: Analysis of Investigational New Drug Applications in 2015. J Clin Oncol 2017;35:3745–52. doi:10.1200/JCO.2017.73.4186

8 Kim ES, Bruinooge SS, Roberts S, et al. Broadening Eligibility Criteria to Make Clinical Trials More Representative: American Society of Clinical Oncology and Friends of Cancer Research Joint Research Statement. J Clin Oncol 2017;35:3737–44. doi:10.1200/JCO.2017.73.7916

9 Scher KS, Hurria A. Under-Representation of Older Adults in Cancer Registration Trials: Known Problem, Little Progress. J Clin Oncol 2012;30:2036–8. doi:10.1200/JCO.2012.41.6727

10 Pavis S, Morris AD. Unleashing the power of administrative health data: the Scottish model. Public Health Res Pr 2015;25. doi:10.17061/phrp2541541

11 Künn S. The challenges of linking survey and administrative data. IZA World of Labor 2015;214. doi:10.15185/izawol.214.

12 Harron K, Dibben C, Boyd J, et al. Challenges in administrative data linkage for research. Big Data & Society 2017;4:205395171774567. doi:10.1177/2053951717745678

13 Oken MM, Creech RH, Tormey DC, et al. Toxicity and response criteria of the Eastern Cooperative Oncology Group. Am J Clin Oncol 1982;5:649–56.

14 Scottish Government. Beating cancer: ambition and action. Edinburgh, UK: Scottish Government 2016. https://www.gov.scot/publications/beating-cancer-ambition-action/ (accessed 15 Oct 2020).

15 Scottish Government. eHealth Strategy 2014-2017. Edinburgh, UK: Scottish Government 2015. https://www.gov.scot/publications/ehealth-strategy-2014-2017/ (accessed 15 Oct 2020).

16 NHS Scotland. West of Scotland Cancer Network. West of Scotland Cancer Network (WoSCAN). https://www.woscan.scot.nhs.uk/ (accessed 10 Dec 2020).

17 NHS Scotland. SCAN Scotland – Regional Cancer Network. South East Scotland Cancer Network (SCAN). https://www.scan.scot.nhs.uk/ (accessed 10 Dec 2020).

18 NHS Scotland. North Cancer Alliance. North Cancer Alliance (NCA). https://www.nhsscotlandnorth.scot/nca (accessed 10 Dec 2020).

19 CIS Oncology. ChemoCare goes live across NHS Scotland. https://www.cis-healthcare.com/latest/chemocare-goes-live-across-nhs-scotland/ (accessed 15 Oct 2020).

20 Information Services Division. Data Dictionary A-Z: CHI number. ISD Scotland Data Dictionary. https://www.ndc.scot.nhs.uk/Dictionary-A-Z/Definitions/index.asp?ID=128&Title=CHI%20Number (accessed 15 Oct 2020).

21 Public Health Scotland. Public Benefit and Privacy Panel for Health and Social Care -HSC-PBPP. 2020. https://www.informationgovernance.scot.nhs.uk/pbpphsc/ (accessed 15 Oct 2020).

22 NHS Greater Glasgow & Clyde. Glasgow Safe Haven. 2020. https://www.nhsggc.org.uk/about-us/professional-support-sites/safe-haven/ (accessed 15 Oct 2020).

23 Baillie K, Mueller T, Pan J, et al. Use of record linkage to evaluate treatment outcomes and trial eligibility in a real-world metastatic prostate cancer population in Scotland. Pharmacoepidemiol Drug Saf 2020;29:653–63. doi:10.1002/pds.4998

24 Scottish Government. Defining Scotland by Rurality. Scottish Government Urban Rural Classification. 2020. https://www2.gov.scot/Topics/Statistics/About/Methodology/UrbanRuralClassification (accessed 15 Oct 2020).

25 Scottish Government. The Scottish Index of Multiple Deprivation. Statistics. 2020. https://www2.gov.scot/SIMD (accessed 15 Oct 2020).

26 Union for International Cancer Control. What is TNM? Resources. 2020. https://www.uicc.org/resources/tnm (accessed 18 Nov 2020).

27 FIGO Committee on Gynecologic Oncology. FIGO staging for carcinoma of the vulva, cervix, and corpus uteri. Int J Gynaecol Obstet 2014;125:97–8. doi:10.1016/j.ijgo.2014.02.003

28 Villaruz LC, Socinski MA. The Clinical Viewpoint: Definitions, Limitations of RECIST, Practical Considerations of Measurement. Clin Cancer Res 2013;19:2629–36. doi:10.1158/1078-0432.CCR-12-2935

29 Zhu J, Yang Y, Tao J, et al. Association of progression-free or event-free survival with overall survival in diffuse large B-cell lymphoma after immunochemotherapy: a systematic review. Leukemia 2020;34:2576–91. doi:10.1038/s41375-020-0963-1

30 Tate JR. The Paraprotein – an Enduring Biomarker. Clin Biochem Rev 2019;40:5–22.

31 Cai T, Giannopoulos AA, Yu S, et al. Natural Language Processing Technologies in Radiology Research and Clinical Applications. Radiographics 2016;36:176–91. doi:10.1148/rg.2016150080

32 McTaggart S, Nangle C, Caldwell J, et al. Use of text-mining methods to improve efficiency in the calculation of drug exposure to support pharmacoepidemiology studies. Int J Epidemiol 2018;47:617–24. doi:10.1093/ije/dyx264

33 Greenfield S, Platt R. Can Observational Studies Approximate RCTs? Value Health 2012;15:215– 6. doi:10.1016/j.jval.2012.01.003

34 Greenfield S. Making Real-World Evidence More Useful for Decision Making. Value Health 2017;20:1023–4. doi:10.1016/j.jval.2017.08.3012

35 Scottish Government. Recovery and Redesign: An Action Plan for Cancer Services. Edinburgh, UK: 2020. https://www.gov.scot/publications/recovery-redesign-action-plan-cancer-services/ (accessed 12 Jan 2021).

36 Mueller T, Alvarez-Madrazo S, Robertson C, et al. Comparative safety and effectiveness of direct oral anticoagulants in patients with atrial fibrillation in clinical practice in Scotland. Br J Clin Pharmacol 2019;85:422–31. doi:10.1111/bcp.13814

37 Fitton CA, Fleming M, Steiner MFC, et al. In Utero Antihypertensive Medication Exposure and Neonatal Outcomes. Hypertension 2020;75:628–33. doi:10.1161/HYPERTENSIONAHA.119.13802

38 Robertson C, Pan J, Kavanagh K, et al. Cost burden of Clostridioides difficile infection to the health service: A retrospective cohort study in Scotland. J Hosp Infect Published Online First: 24 July 2020. doi:10.1016/j.jhin.2020.07.019

39 Chou W-Y, Lai P-Y, Hu J-M, et al. Association between atopic dermatitis and colorectal cancer risk: A nationwide cohort study. Medicine 2020;99. doi:10.1097/MD.0000000000018530

40 Ong ELH, Goldacre R, Hoang U, et al. Subsequent Primary Malignancies in Patients with Nonmelanoma Skin Cancer in England: A National Record-Linkage Study. Cancer Epidemiol Biomarkers Prev 2014;23:490–8. doi:10.1158/1055-9965.EPI-13-0902

41 Houben E, van Haalen HGM, Sparreboom W, et al. Chemotherapy for ovarian cancer in the Netherlands: a population-based study on treatment patterns and outcomes. Med Oncol 2017;34:50. doi:10.1007/s12032-017-0901-x

42 Kurian AW, Lichtensztajn DY, Keegan THM, et al. Patterns and predictors of breast cancer chemotherapy use in Kaiser Permanente Northern California, 2004-2007. Breast Cancer Res Treat 2013;137:247–60. doi:10.1007/s10549-012-2329-5

43 Khozin S, Carson KR, Zhi J, et al. Real-World Outcomes of Patients with Metastatic Non-Small Cell Lung Cancer Treated with Programmed Cell Death Protein 1 Inhibitors in the Year Following U.S. Regulatory Approval. Oncologist 2019;24:648–56. doi:10.1634/theoncologist.2018-0307

44 Pezzi CM. Big Data and Clinical Research in Oncology: The Good, the Bad, the Challenges, and the Opportunities. Ann Surg Oncol 2014;21:1506–7. doi:10.1245/s10434-014-3519-7

45 Weber SC, Seto T, Olson C, et al. Oncoshare: Lessons Learned from Building an Integrated Multi-institutional Database for Comparative Effectiveness Research. AMIA Annu Symp Proc 2012;2012:970–8.

46 van Herk-Sukel MPP, van de Poll-Franse LV, Lemmens VEPP, et al. New opportunities for drug outcomes research in cancer patients: the linkage of the Eindhoven Cancer Registry and the PHARMO Record Linkage System. Eur J Cancer 2010;46:395–404. doi:10.1016/j.ejca.2009.09.010

47 Wiitala WL, Vincent BM, Burns JA, et al. Variation in laboratory naming conventions in EHRs within and between hospitals: A nationwide longitudinal study. Med Care 2019;57:e22–7. doi:10.1097/MLR.0000000000000996

48 Clauser SB, Wagner EH, Bowles EJA, et al. Improving Modern Cancer Care Through Information Technology. Am J Prev Med 2011;40:S198–207. doi:10.1016/j.amepre.2011.01.014

49 Kanas G, Morimoto L, Mowat F, et al. Use of electronic medical records in oncology outcomes research. Clinicoecon Outcomes Res 2010;2:1–14.

50 Scottish Government. Medicines Use and Digital Capabilities - Building capability to assess real-world benefits, risks and value of medicines: Towards a Scottish Medicines Intelligence Unit. Data Scoping Taskforce 2018. https://www2.gov.scot/Resource/0054/00540468.pdf (accessed 15 Oct 2020).

51 Scottish Government. A Charter for Safe Havens in Scotland. Edinburgh, UK: Scottish Government 2015. https://www.gov.scot/binaries/content/documents/govscot/publications/agreement/2015/11/charter-safe-havens-scotland-handling-unconsented-data-national-health-service-patient-records-support-research-statistics/documents/charter-safe-havens-scotland-handling-unconsented-data-national-health-service-patient-records-support-research-statistics/charter-safe-havens-scotland-handling-unconsented-data-national-health-service-patient-records-support-research-statistics/govscot%3Adocument/00489000.pdf (accessed 15 Oct 2020).

52 Hanna C, Lemmon E, Ennis H, et al. Creation of the first national linked colorectal cancer dataset in Scotland: prospects for future research and a reflection on lessons learned [t: Creation of Scottish colorectal cancer dataset for research purposes. IJPDS 2021;6. doi:10.23889/ijpds.v6i1.1654

53 Information Services Division. National Data Catalogue: National Datasets. ISD Scotland National Data Catalogue. https://www.ndc.scot.nhs.uk/National-Datasets/index.asp (accessed 15 Oct 2020).

54 NHS National Services Scotland. SCI store. Scottish Care Information. https://www.sci.scot.nhs.uk/products/store/store_main.htm (accessed 15 Oct 2020).

